# Population Pharmacokinetic Modeling of Intravenous Topiramate in Patients with Epilepsy and Migraine

**DOI:** 10.64898/2026.02.26.26346744

**Authors:** Adeboye O. Bamgboye, Lisa D. Coles, Bovornpat Suriyapakorn, Usha Mishra, Robert L. Kriel, Ilo E. Leppik, James R. White, James C. Cloyd

## Abstract

Topiramate (TPM) is approved for seizures and migraine prophylaxis and is used off-label for several neuropsychiatric conditions. The available dosage forms, including tablets and sprinkle capsules, are unsuitable for patients who may be unable to take medicine orally. The resulting potential treatment interruptions could have untoward consequences and underscores the importance of developing a parenteral formulation. In this study, we developed a population pharmacokinetic model of a novel, intravenous TPM formulation using data from a study in patients with epilepsy or migraine receiving a single intravenous dose of stable-labeled TPM.

In total, 246 TPM concentrations from 20 adult patients were included for model development. A three-compartment pharmacokinetic model with linear elimination fit the concentration-time data best. Simulations for various loading and maintenance regimens for patients with and without enzyme-inducing comedications were performed.

The final estimates(95% confidence interval (CI)) for CL (L/h), V1 (L), and the peripheral volumes, V2 and V3 for a 70 kg person were 1.31(**1.01 – 1.53**), 9.84 (**8.49 – 11.0**), 39.1 (**36.5 – 41.8**)L, and 9.01 (**6.41 – 44.3**) respectively. The use of enzyme-inducing co-medication was the only significant covariate, associated with a 63% increase in clearance .Goodness-of-fit plots and visual predictive checks indicate satisfactory model performance and prediction. The simulation results indicate that adjusting doses for patients receiving IV TPM can mitigate the changes in plasma TPM concentrations resulting from enzyme induction. This population pharmacokinetic model for intravenous topiramate can inform dosing decisions for patients with epilepsy when used as either initiation or bridging therapy.

## INTRODUCTION

Topiramate (TPM) is approved for seizures and migraine prophylaxis and is used off-label for a growing list of neuropsychiatric conditions ^1, 2^. Currently approved TPM formulations include 25 to 200 mg tablets and sprinkle capsules. However, patients who cannot take oral TPM face potential treatment interruptions with untoward consequences. Development of an intravenous topiramate formulation is underway, and some studies have been published reporting the safety and pharmacokinetics in healthy individuals and patients ^3^, ^4^, ^5^

Topiramate pharmacology has been extensively studied. Mechanisms of action include blocking voltage-gated Na^+^ ^6^ and Ca^2+^ channels ^7^, ^8^ enhancing GABA-mediated transmission ^9^, ^10^, glutamate-mediated transmission ^11, 12^, and inhibiting the carbonic anhydrase enzyme ^13, 14, 15^. However, the role of these mechanisms in terms of its anti-seizure effect is not well understood.

TPM exhibits linear pharmacokinetics at clinical doses, rapid absorption, and a half-life of 20-30 hours. ^2, 16, 3^ Approximately 70-80% of an administered dose is excreted unchanged in the urine ^17^. Several population pharmacokinetic models of oral TPM have been developed to characterize its pharmacokinetics and identify covariates that influence the pharmacokinetic parameters ^18, 19, 20, 21^. Although one study characterized the population pharmacokinetics of intravenous topiramate in healthy individuals ^21^, to our knowledge, there is no published population pharmacokinetic model of intravenous TPM in patients with epilepsy or migraine.

In this study, we developed a population pharmacokinetic model of a novel, intravenous TPM formulation using data from a pilot phase I study in patients receiving a single intravenous dose of stable-labeled TPM ^4^. A portion of those patients were taking enzyme-inducing comedications. The stable-labeled TPM formulation includes a ^13^C-labeled (nonradioactive) isotope, which allows for safe and rigorous characterization of the pharmacokinetics of IV TPM without the need for washout or interruption of oral TPM therapy in these patients.^22^ The objectives of this study were to: (i) estimate the population pharmacokinetic parameters of IV TPM and their interindividual variability, (ii) identify potential covariates including enzyme-inducing comedications, influencing the estimated parameters, and (iii) simulate various loading and maintenance regimens for patients with and without inducing comedications.

## METHODS

### Study Population

Twenty adult patients receiving maintenance oral TPM therapy for epilepsy or migraine and administered a single 25mg intravenous dose of stable-labeled IV TPM, were included in this analysis. Seven (35%) were receiving enzyme-inducing comedications. Of these, 4 were on carbamazepine, 1 on phenytoin, 1 on oxcarbazepine, and 1 was taking both carbamazepine and phenytoin. Individuals who were pregnant or breastfeeding were excluded. A history of intolerance to intravenous medication administration and known hypersensitivity to TPM were additional exclusion criteria for the study. Information about age, race, sex, renal function, and medical history were collected at the screening visit. Renal function was estimated using the Cockcroft-Gault equation (CrCL). ^23^

### Study drug formulation and measurement

The study drug was formulated as a 10mg/mL solution dissolved in a 10% sulfobutyl cyclodextrin (Captisol®, Ligand Technologies, La Jolla, CA, USA). Plasma concentrations of the stable-labeled IV TPM and oral TPM were quantified using a validated LC-MS method with electrospray ionization and selected ion monitoring, utilizing distinct m/z transitions (m/z 338, m/z 344, and m/z 350 for oral TPM, stable-labeled IV TPM, and internal standard respectively) to enable separate analyte quantification. A full description of the analytical method and study drug formulation is provided in a previous publication ^4^.

### Study design

The study was conducted under IND #78993 and approved by the University of Minnesota Institutional Review Board. All participants provided written informed consent.

A 25 mg dose of stable-labeled IV TPM was administered to patients along with their usual morning oral TPM dose. The infusion was administered over 10 minutes under monitoring conditions. Blood samples were drawn at predose, 5, 15, and 30 min, and 1, 2, 4,6, 12, 24, 48, 72, and 96 hours after administration. Measurements of sodium, potassium, glucose, serum creatinine, urea nitrogen, hemoglobin, liver enzymes, and total plasma protein were done prior to IV administration. A full description of the study design is provided in a previous publication. ^4^

### Population pharmacokinetic (PopPK) model development

The PopPK model was developed using nonlinear mixed-effects modelling software NONMEM (version 7.5.1; ICON Development Solutions, Ellicott City, MD, USA) with a Finch Studio interface (Enhanced Pharmacodynamics, LLC). The first-order conditional estimation with interaction (FOCEI) was used for estimation of parameters. Model selection was based on a significant change in the objective function value (OFV), Akaike information criterion (AIC), visual inspection of goodness-of-fit (GOF) plots, and visual predictive check. One-, two- and three-compartment PK models with linear elimination were tested. The models were parameterized using volume and clearance terms. The base model was designed to incorporate the effect of weight with a fixed allometric exponent of 0.75 for clearances and 1 for the volumes to balance model complexity with physiological plausibility. The inter-individual variability (IIV) was estimated using an exponential random effect model, consistent with standard PopPK modeling practice and ensuring parameter positivity, as described by the following equation:

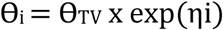

Where ***ϴi*** is the individual PK parameter estimate, ***ϴTV*** is the typical value of the PK parameter, and ***ηi*** is the interindividual random effect with mean of 0 and variance of ⍵^2^. The estimates of the IIV are presented as coefficients of variation expressed as a percentage (%CV) calculated using the equation below:

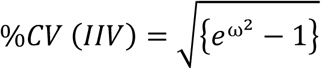

For the residual error model, we tested additive, proportional and combined residual error models. The base model was selected based on the likelihood ratio test (LRT) of change in the OFV. Covariate analysis was conducted using a stepwise method based on the changes in OFV using a LRT to evaluate the factors influencing TPM PK. The covariates tested included age, height, sex, inducer status, and creatinine clearance. These were added to the base model using a forward inclusion criterion of p < 0.05(χ^2^, df = 1, **Δ**OFV of at least 3.84) and a backward elimination criterion of p < 0.01(χ^2^, df =1, **Δ**OFV of at least 6.63) respectively.

### Model evaluation

Model adequacy was assessed by successful minimization, significant changes in the objective function value (OFV) and Akaike information criterion (AIC), visual inspection of goodness-of-fit (GOF) plots and visual predictive check, precision of parameter estimates, and bootstrap resampling (n =1000).

### Model simulation

We performed simulations to evaluate various IV TPM maintenance dosing regimens in virtual patients (n = 200 each) with and without concomitant enzyme-inducing medications. We also performed loading dose simulations in a reference individual (70kg, without enzyme induction) to evaluate concentration-time profiles across alternative loading doses.

## RESULTS

### Patient characteristics

A total of 246 PK observations (unique TPM concentrations) from 20 patients (13 females, 7 males, all Caucasian) were included in the analysis. The mean (SD) age for the patients was 39.8(12.1) years, and the median(range) weight was 85.2(54.5–150.3) kg. The median creatinine clearance was 107.6 mL/min (range: 46–206). The baseline characteristics of the studied population are summarized in **Table 1**. The concentration-time profiles (0–96 hours) following a 25mg intravenous dose stratified by inducer status is shown in **Figure 1**.

**Table 1.** Demographic and baseline characteristics.

| Parameters | Patients |
| --- | --- |
| n | 20 |
| Age(years) |  |
| *Mean (SD)* | 39.8 (12.1) |
| *Median(range)* | 41.5 (26 - 74) |
| Height(cm) |  |
| *Mean (SD)* | 169 (8.36) |
| *Median(range)* | 170 (155 – 182) |
| Weight (Kg) |  |
| *Mean (SD)* | 92.3 (26.6) |
| *Median(range)* | 85.2 (54.5 – 150.3) |
| Sex, n (%) |  |
| *Female* | 13 (65) |
| *Male* | 7 (35) |
| Race, n (%) |  |
| *White* | 20 (100) |
| Creatinine Clearance(mL/min) |  |
| *Mean (SD)* | 106.7 (34.69) |
| *Median(range)* | 102.5 (46 – 206) |
| Inducer comedications, n (%) |  |
| *Received enzyme inducer* | 7 (35) |
| *No enzyme inducer* | 13 (65) |

**Figure 1.**
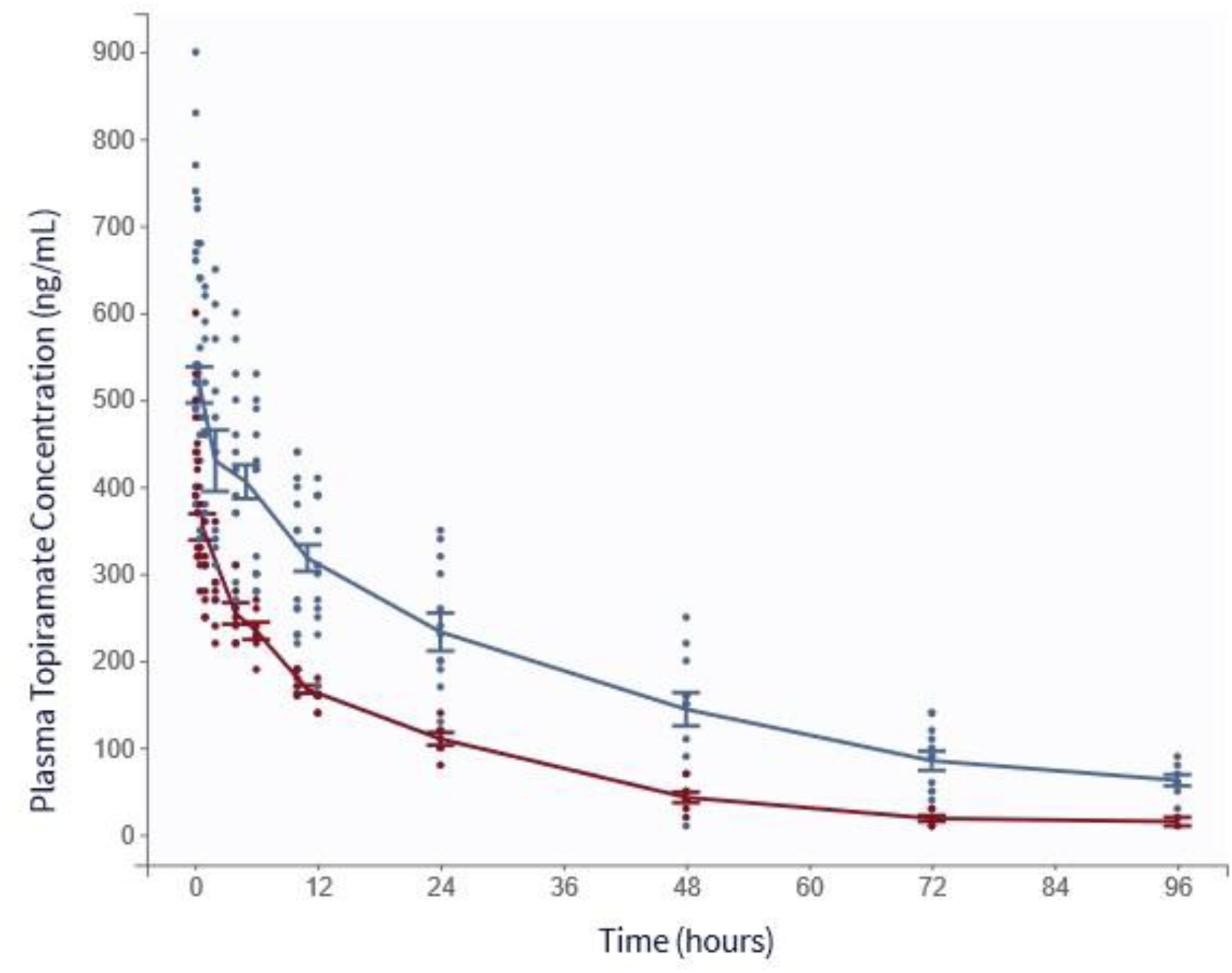
Mean concentration-time profile for patients after intravenous topiramate (0–96 hours). Blue line represents profile for individuals on monotherapy. Red line represents profile for patients receiving enzyme-inducing comedications. Points represents observations. Error bars represent standard errors.

### Population PK Model

The final model was a three-compartment model with linear elimination and an exponential IIV model for central clearance and volume. The final population PK parameters are presented in **Table 2**. Correlation parameters between CL, V1 and V2, and between V1 and V2 improved the model fit and were subsequently included. A proportional error model best described the residual unexplained variability. Results of the structural model selection and empirical Bayes estimate reliability assessment are presented in **Tables S1** and **S2**, respectively. During the forward inclusion, Inducer on CL was the only significant covariate (ΔOFV > 3.84). Backward elimination resulted in a significant rise in OFV (ΔOFV > 6.63); therefore, the final model included the influence of inducer status on CL. The results of the covariate analysis are summarized in **Table S3**. The quantitative relationships between model parameters and covariates are presented below:

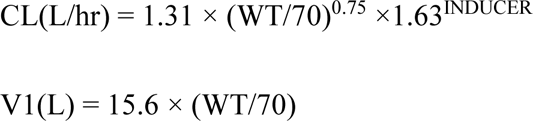

**Table 2.** Parameter estimates of the final pharmacokinetic model.

| Final Model |  | Bootstrap Model (n = 1000) |  |  |  |
| --- | --- | --- | --- | --- | --- |
| Parameter | Fixed effects | RSE | Bootstrap | 95% CI | Shrinkage (%) |
|  | estimate | (%) | Median |  |  |
| **CL (L/hr)** | $1.31 \times (\text{WT}/70)^{0.75}$ | 9.51 | 1.28 | 1.01–1.53 | — |
| **V1 (L)** | $9.84 \times (\text{WT}/70)$ | 8.7 | 9.83 | 8.49–11.0 | — |
| **Q2 (L/hr)** | $197 \times (\text{WT}/70)^{0.75}$ | 6.73 | 199 | 181–223 | — |
| **V2 (L)** | $39.1 \times (\text{WT}/70)$ | 5.1 | 38.9 | 36.5–41.8 | — |
| **Q3 (L/hr)** | $0.6 \times (\text{WT}/70)^{0.75}$ | 41.7 | 0.6 | 0.4–1.2 | — |
| **V3 (L)** | $9.01 \times (\text{WT}/70)$ | 18.8 | 9.44 | 6.41–44.3 | — |
| **Covariate** |  |  |  |  |  |
| **Inducer ~ CL** | 1.63 | 16.3 | 1.7 | 1.12–2.30 | — |
| **Interindividual variability** |  |  |  |  |  |
| **IIV on CL (CV%)** | 33.5 | 40 | 32.4 | 20.2–52.6 | 1.48 |
| Correlation ($\Omega_{\text{CL}, \text{V1}}$) | 0.06 | 51.7 | 0.05 | 0.01–0.12 | — |
| **IIV on V1 (CV%)** | 18.1 | 74.2 | 17.5 | 5.5–32.0 | 1.5 |
| Correlation ($\Omega_{\text{V1}, \text{V2}}$) | 0.03 | 52.8 | 0.02 | 0.005–0.05 | — |
| Correlation ($\Omega_{\text{CL}, \text{V2}}$) | 0.03 | 60.4 | 0.03 | 0.005–0.07 | — |
| **IIV on V2 (CV%)** | 19.3 | 37.6 | 17.5 | 11.9–24.1 | 4.21 |
| **Residual variability** |  |  |  |  |  |
| **Proportional (%CV)** | 0.02(12.8) | 13.7 | 0.02 | 0.01–0.02 | 6.28 |
| CL: central clearance; V1: central volume of distribution; Q2: intercompartmental clearance of the first peripheral compartment; V2: volume of distribution of the first peripheral compartment; Q3: intercompartmental clearance of the second peripheral compartment; V3: volume of distribution of the second peripheral compartment; RSE: relative standard error; WT: bodyweight; CV: coefficient of variation $\%CV = \sqrt{(e^{\omega^2} - 1)}$; IIV: interindividual variability | | | | | |

### Model evaluation

The GOF plots are shown in **Figure 2**. Visual inspection of the GOF plots show that the final model adequately captures the trend in the data. The observations (DV) versus population predictions (PRED) and individual predictions (IPRED) plots show a symmetrical distribution around the diagonal line indicating good model fit. The CWRES vs PRED and Time plots show that a majority of residuals were within two standard deviations and evenly distributed around the line of identity indicating proper specification of the structural and residual models. The means and 95% confidence intervals of the final PK parameters are shown in **Table 2**. Bootstrap resampling with 1000 replicates was implemented. The medians of the PK parameters from bootstrap analysis were similar to the model estimates indicating robustness of the parameter estimates. The VPC plot (**Figure 3)** shows the model’s reliability in capturing the trend and variability in the data set.

**Figure 2.**
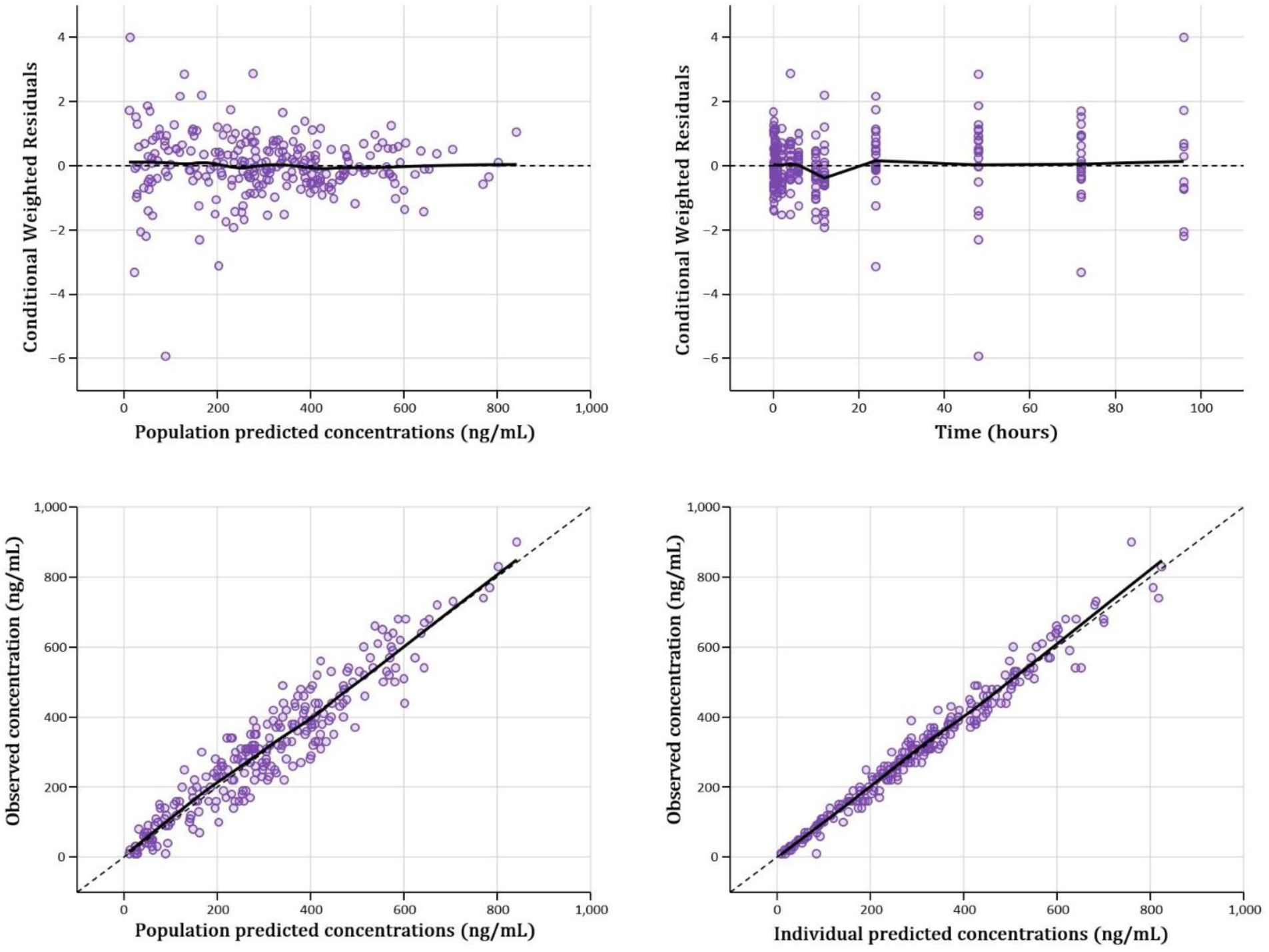
Goodness-of-fit plots of the final model. Conditional weighted residuals (CWRES) versus population predictions plot (upper left), CWRES versus time plot (upper right), individual observations (DV) versus population predicted concentrations (PRED) plot (lower left), and DV versus individual predicted concentrations (IPRED) plot (lower right).

**Figure 3.**
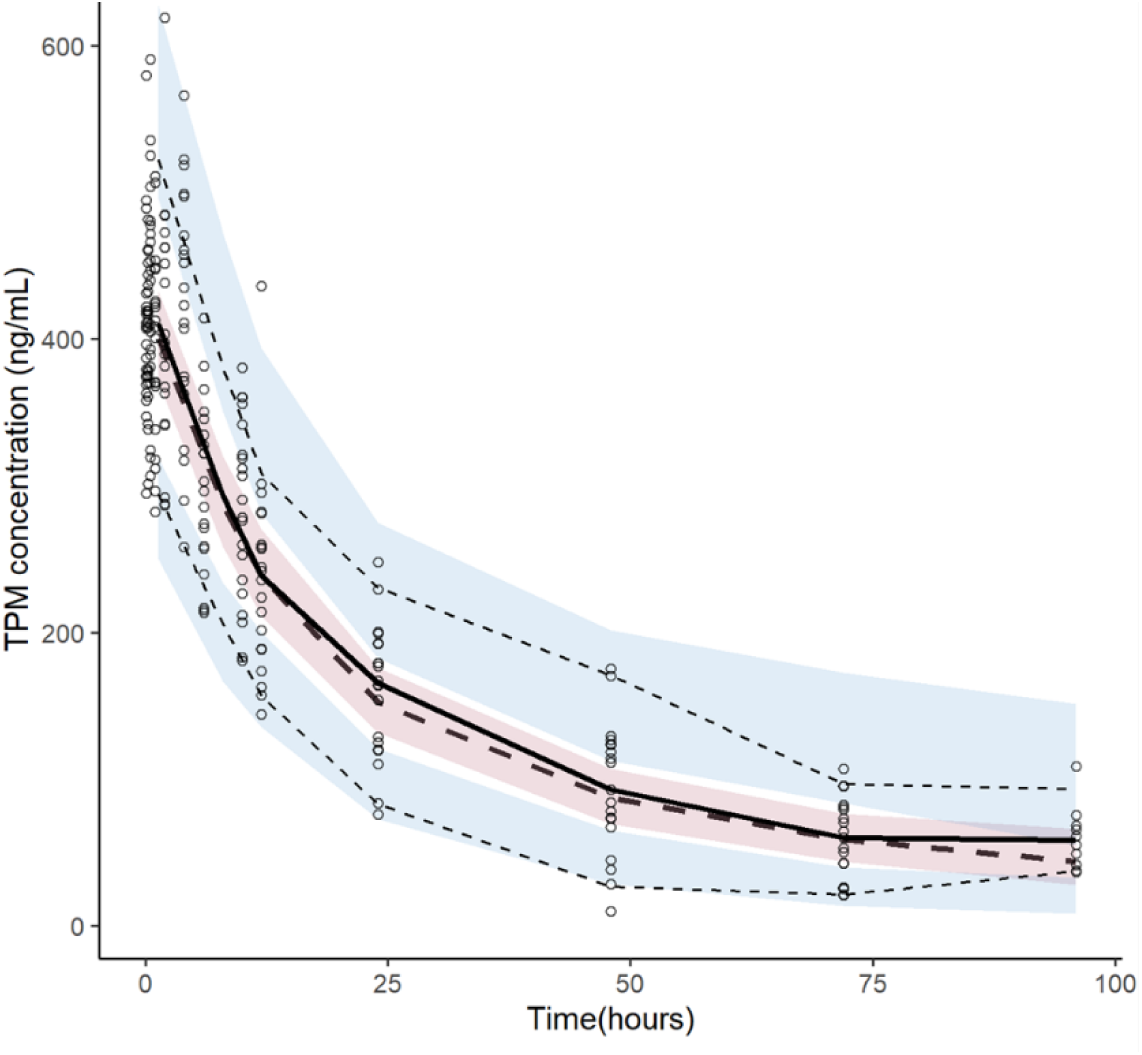
Visual predictive check of observed intravenous TPM concentrations. Open circles represent observed data. Solid line represents the median of observed concentrations. The dashed black line in the middle represents the median of the simulated concentrations. The upper and lower dashed lines represent the observed 5th and 95th percentiles respectively. Shaded pink represents the confidence interval around simulated median. Shaded blue areas represent the confidence interval around the 5th and 95th percentiles of the simulated data.

### Model simulation

The loading dose simulations showed dose-proportional increases in Cmax across doses irrespective of enzyme induction status (**Figure 4**). The maintenance dose simulations showed differences in the concentration-time profiles between inducer and non-inducer groups at 50 and 100 mg TPM doses (**Figures 5a and 5c**). Dose adjustments for patients on enzyme-inducing comedications led to comparable profiles with patients not on enzyme-inducing comedications (**Figures 5b and 5d**).

**Figure 4.**
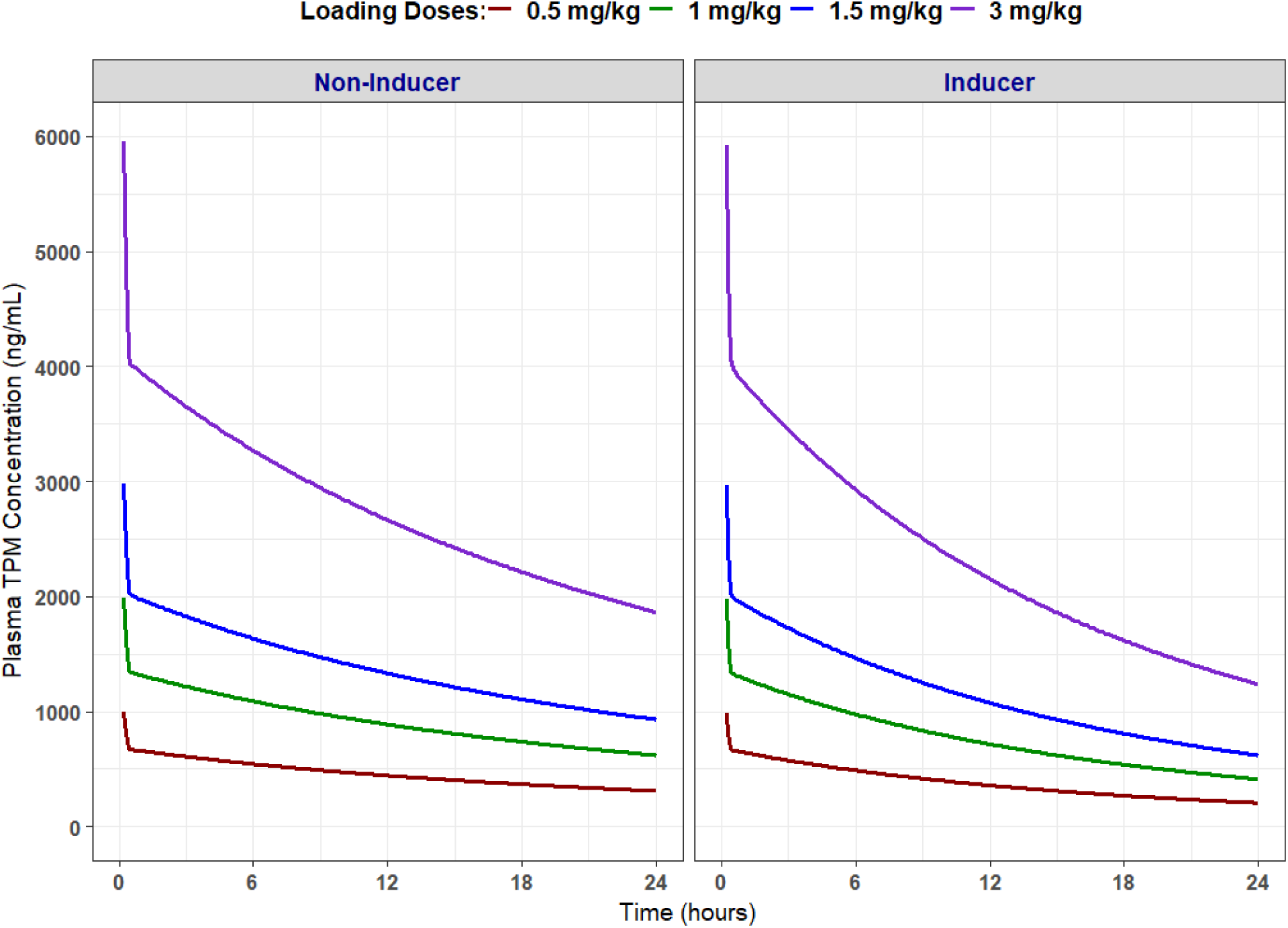
Simulated plasma concentration-time profiles following various loading doses of intravenous TPM in a 70kg individual with and without concurrent enzyme inducers.

**Figure 5.**
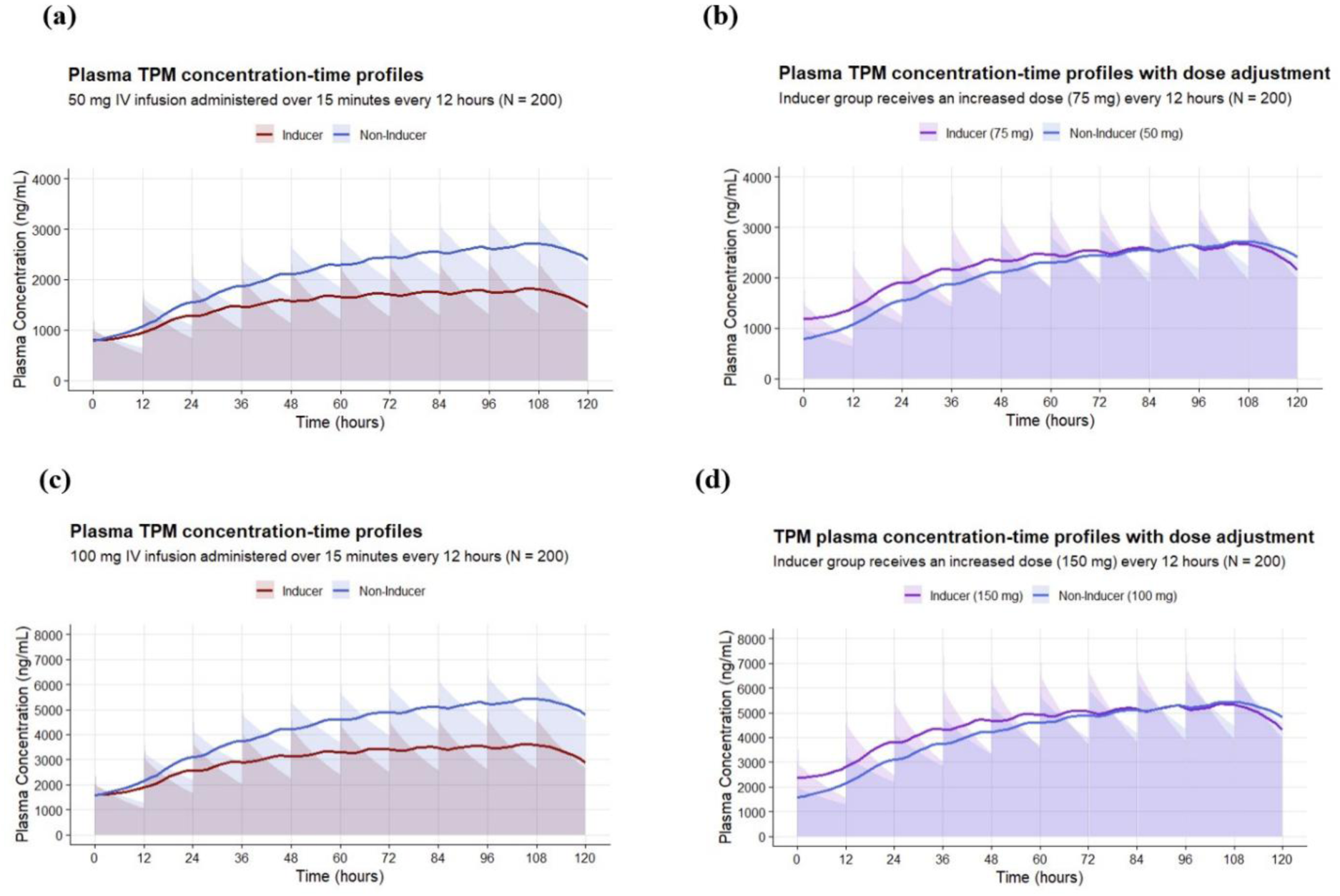
Simulated plasma concentration-time profiles for two intravenous topiramate (TPM) maintenance regimens in patients with and without enzyme-inducing comedications. TPM doses were administered over a 15-minute period every 12 hours. Solid lines represent the mean concentration for each group. (a) 50 mg IV dose; (b) adjusted dose (75 mg IV); (c) 100 mg IV dose; (d) adjusted dose(150 mg IV).

## DISCUSSION

In this study we found a three-compartment pharmacokinetic model fit the TPM concentration-time data best for patients with epilepsy and migraines. We also found that enzyme-inducing comedications significantly increased TPM clearance in this population. In contrast to many other population pharmacokinetic models of oral topiramate that were described by a one- or two-compartment model, a three-compartment model fit our data best. While many studies with oral topiramate used sparse steady-state concentrations for modeling, our study used rich sampling, and we believe this approach enabled better characterization of TPM PK. Notably, another PopPK study of IV and oral TPM with rich sampling in healthy volunteers supported a two-compartmental structural model ^21^. In that study, the authors concurrently modeled oral and IV TPM concentration data. In our model, topiramate central clearance for an adult (70kg) not on inducing comedications was estimated to be 1.31 L/hr which is consistent with clearance estimates (1.16 – 1.47 L/hr) from previous studies of TPM in healthy volunteers and patients with epilepsy ^17, 3, 19^.

We found that TPM clearance increased by 63% (coefficient for inducer effect = 1.63) in patients receiving enzyme inducers. This finding is consistent with previously published studies reporting increases of 15-94% ^24, 20, 17^. A direct consequence of the increased clearance is the need to adjust doses for patients receiving TPM with enzyme-inducing comedications. While some studies modeled the influence of single enzyme-inducing comedications ^19, 17^, data limitations and a need to avoid overparameterization led to a simplified binary covariate approach for modeling enzyme inducers’ effect on clearance. Our PopPK simulations show a notable difference in the concentration-time profiles for patients receiving 50 and 100 mg doses of TPM while on and off enzyme-inducers (**Figure 5**). Increasing the doses for patients on enzyme-inducing comedications led to comparable profiles with patients not on enzyme-inducing comedications (**Figure 5**). While dose modifications may be recommended for patients receiving enzyme-inducing comedications, changes to loading doses would not be warranted in this population since loading doses are typically dependent on volume of distribution which is not impacted by enzyme-inducers (**Figure 4**).

The observed increase in clearance is notable and raises important questions about the mechanisms that may contribute to this increase. Given that TPM is primarily excreted renally, the considerable difference in CL suggests that both hepatic and renal CL are impacted by enzyme-inducing medications. Furthermore, it is plausible that enzyme-inducing medications interact with active transporters in the renal tubules that play a role in the renal excretion of TPM. Further studies are needed to adequately describe the extrahepatic mechanisms involved in TPM clearance when co-administered with enzyme inducers.

We also investigated the influence of other covariates on IV TPM clearance. Age, sex, creatinine clearance (CrCL), and height were not significant covariates for clearance. While a previous study found clearance to increase with age ^17^, no such relationship was identified in our study, possibly reflecting age distribution differences between studies.. Similarly, renal function has been reported to influence TPM clearance in several published studies. ^17, 19, 18^ We did not include race in the covariate modeling since we lacked the data to reliably estimate the effect of race. One study ^21^ included race as a covariate but did not find a relationship between clearance and race after accounting for renal function ^21^. At present, there is no evidence to suggest that TPM clearance differs across racial groups; however, further research is needed.

The standard errors of the final model were acceptable and demonstrate reliable estimation of parameters. The interindividual variability of clearance was 33.5% and this is consistent with previous studies ^25^, ^17^, ^20^ demonstrating considerable variability in drug distribution among individuals. Bootstrap and VPC results indicate that the model exhibits stability and satisfactory reproducibility.

It is worth mentioning some limitations of our study. Condensing the enzyme inducer effect into a single covariate assumes that the changes to TPM clearance as a function of enzyme-inducing comedications are uniform across commonly co-administered medications like Carbamazepine and Phenytoin. Results from previous studies suggest that the increase in TPM clearance may depend on the specific inducing co-medication administered ^17, 19^. While previous studies have shown that renal function influences TPM clearance ^17, 19^, ^18^, there was no significant relationship between CrCL and TPM clearance in our model. This lack of an effect could be due to the near-normal CrCL values of the patient population. As a result, our model may not generalize well to patient populations with renal impairment. Future studies evaluating renal function, including CrCL as a covariate for IV TPM clearance in larger patient cohorts are warranted to improve the clinical utility of the model.

## CONCLUSION

In conclusion, we successfully developed a population model to describe intravenous topiramate pharmacokinetics in patients with epilepsy and migraine. The model adequately captured the data trend as seen from the standard error estimates of the parameters and visual inspection of the diagnostic plots. Inducer status was the only significant covariate that influenced topiramate clearance. IV TPM clearance increases by 63% for individuals receiving concomitant enzyme inducers. Additional studies including patients across a broader range of renal function may improve the precision and clinical utility of the model. Overall, the model provides a useful foundation for characterizing IV TPM characteristics and can inform future dosing decision.

## Acknowledgments

The authors thank Richard Brundage, Thomas Henry, Annie Clark, and Susan Marino who contributed to the design and conduct of the clinical studies that provided data for this population pharmacokinetic analysis.

## Author Contributions

A.O.B., B.S., L.D.C., I.E.L., R.L.K., J.W.R., and J.C.C. wrote the manuscript. L.D.C., J.C.C., and A.O.B., designed the study. U.M., J.C.C., and I.E.L. performed the research. A.O.B., B.S., L.D.C., J.C.C. analyzed the data.

## Data Availability Statement

The data that support the findings of this study are available on request from the corresponding author. The data are not publicly available due to privacy or ethical restrictions.

## Supplementary materials

**Table S1.** Quantitative model selection criteria comparison.

| Model structure | OFV | ΔOFV vs 1-compartment | AIC | BIC | Number of parameters | Comments |
| --- | --- | --- | --- | --- | --- | --- |
| 1-compartment | 2626.2 | – | 2638.2 | 2659.2 | 5 | Baseline |
| 2-compartment | 2153.5 | -472.7 | 2169.5 | 2197.5 | 7 | Considerable improvement |
| 3-compartment | 2113.43 | -512.8 | 2133.4 | 2168.5 | 9 | Significant additional improvement |
| OFV, AIC, and BIC all favor the 3-compartment model |  |  |  |  |  |  |

**OFV:** objective function value, **AIC:** Akaike information criterion, **BIC:** Bayesian information criterion

**Table S2.** Random-effects diagnostics supporting EBE reliability.

| Model | η-shrinkage(%) on CL | η-shrinkage(%) on V1 | η-shrinkage(%) on V2 | ε-shrinkage(%) |
| --- | --- | --- | --- | --- |
| 1-compartment | 4.46 | 4.46 | - | 2.28 |
| 2-compartment | 1.26 | 6.14 | - | 5.33 |
| 3-compartment | 1.48 | 1.50 | 4.21 | 7.55 |

**ETA( $\eta$ ):** interindividual random effects, **EPSILON( $\epsilon$ ):** residual unexplained variability, **EBE:** empirical bayes estimate

**Table S3.**
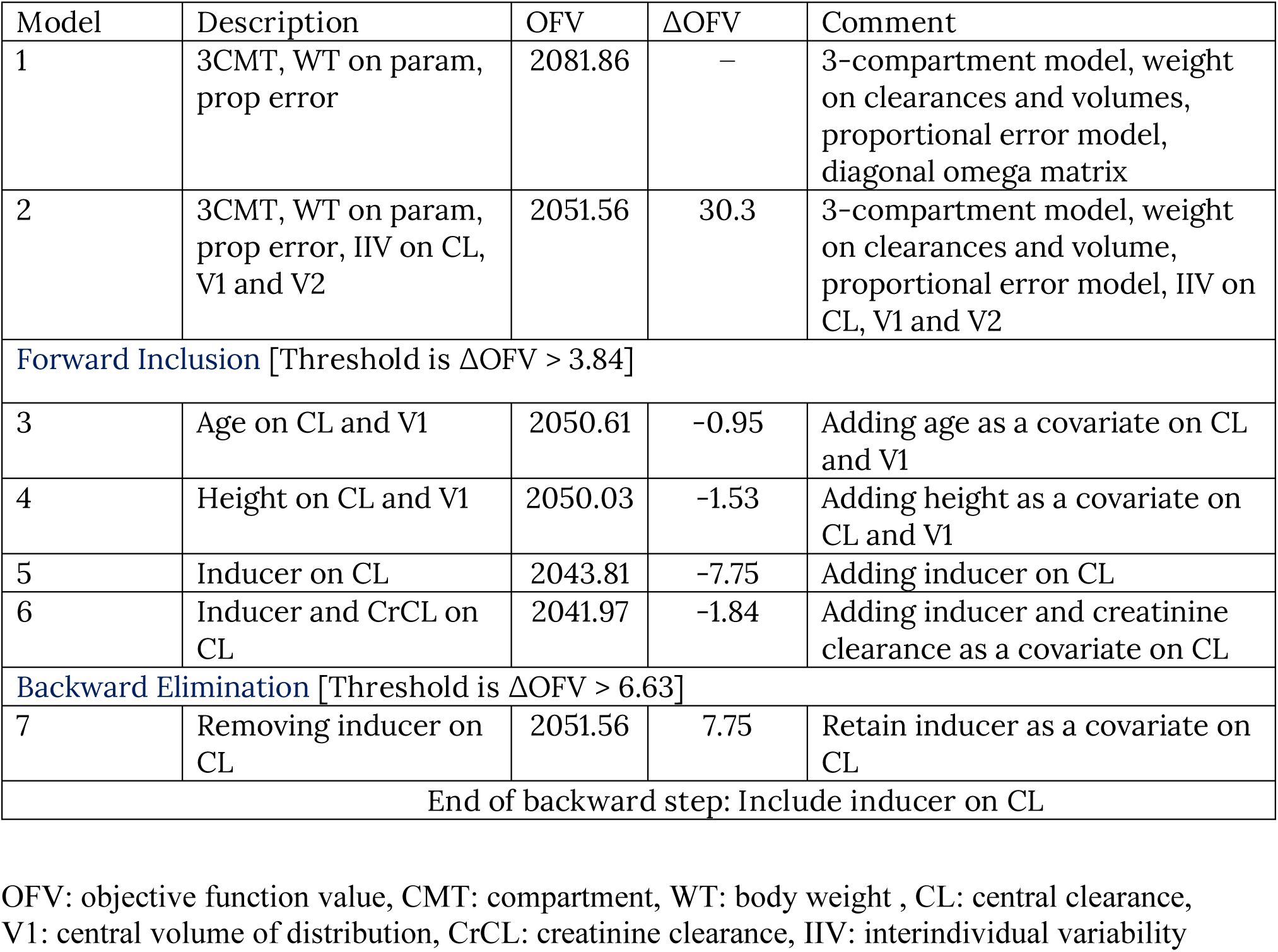
Covariate Search.

## REFERENCES

1. Kanner AM, Bicchi MM. Antiseizure Medications for Adults With Epilepsy: A Review. JAMA. 2022;327(13):1269–1281. doi:10.1001/jama.2022.3880

2. Bai YF, Zeng C, Jia M, Xiao B. Molecular mechanisms of topiramate and its clinical value in epilepsy. Seizure. 2022;98:51–56. doi:10.1016/j.seizure.2022.03.024

3. Clark AM, Kriel RL, Leppik IE, et al. Intravenous topiramate: Comparison of pharmacokinetics and safety with the oral formulation in healthy volunteers. Epilepsia. 2013;54(6):1099–1105. doi:10.1111/epi.12134

4. Clark AM, Kriel RL, Leppik IE, et al. Intravenous topiramate: Safety and pharmacokinetics following a single dose in patients with epilepsy or migraines taking oral topiramate. Epilepsia. 2013;54(6):1106–1111. doi:10.1111/epi.12165

5. Apostolakopoulou L, Bosque Varela P, Rossini F, et al. Intravenous topiramate for seizure emergencies – First in human case report. Epilepsy & Behavior. 2023;142:109158. doi:10.1016/j.yebeh.2023.109158

6. Lin CH, Ho CJ, Lu YT, Tsai MH. Response to Sodium Channel blocking Antiseizure medications and coding polymorphisms of Sodium Channel genes in Taiwanese epilepsy patients. BMC Neurology. 2021;21(1):367. doi:10.1186/s12883-021-02395-2

7. Zhang X lei, Velumian AA, Jones OT, Carlen PL. Modulation of High-Voltage–Activated Calcium Channels in Dentate Granule Cells by Topiramate. Epilepsia. 2000;41(s1):52–60. doi:10.1111/j.1528-1157.2000.tb02173.x

8. Kuzmiski JB, Barr W, Zamponi GW, MacVicar BA. Topiramate Inhibits the Initiation of Plateau Potentials in CA1 Neurons by Depressing R-type Calcium Channels. Epilepsia. 2005;46(4):481–489. doi:10.1111/j.0013-9580.2005.35304.x

9. White HS, Brown SD, Woodhead JH, Skeen GA, Wolf HH. Topiramate modulates GABA-evoked currents in murine cortical neurons by a nonbenzodiazepine mechanism. Epilepsia. 2000;41(S1):17–20.

10. Simeone TA, Wilcox KS, White HS. Subunit selectivity of topiramate modulation of heteromeric GABAA receptors. Neuropharmacology. 2006;50(7):845–857. doi:10.1016/j.neuropharm.2005.12.006

11. Angehagen M, Rönnbäck L, Hansson E, Ben-Menachem E. Topiramate reduces AMPA-induced Ca(2+) transients and inhibits GluR1 subunit phosphorylation in astrocytes from primary cultures. J Neurochem. 2005;94(4):1124–1130. doi:10.1111/j.1471-4159.2005.03259.x

12. Andreou AP, Goadsby PJ. Topiramate in the treatment of migraine: a kainate (glutamate) receptor antagonist within the trigeminothalamic pathway. Cephalalgia. 2011;31(13):1343–1358. doi:10.1177/0333102411418259

13. Dodgson SJ, Shank RP, Maryanoff BE. Topiramate as an inhibitor of carbonic anhydrase isoenzymes. Epilepsia. 2000;41(S1):35–39. doi:10.1111/j.1528-1157.2000.tb06047.x

14. Vullo D, Voipio J, Innocenti A, et al. Carbonic anhydrase inhibitors. Inhibition of the human cytosolic isozyme VII with aromatic and heterocyclic sulfonamides. Bioorg Med Chem Lett. 2005;15(4):971–976. doi:10.1016/j.bmcl.2004.12.052

15. Supuran CT. Carbonic anhydrases: novel therapeutic applications for inhibitors and activators. Nat Rev Drug Discov. 2008;7(2):168–181. doi:10.1038/nrd2467

16. Girgis IG, Nandy P, Nye JS, et al. Pharmacokinetic–pharmacodynamic assessment of topiramate dosing regimens for children with epilepsy 2 to &10 years of age. Epilepsia. 2010;51(10):1954–1962. doi:10.1111/j.1528-1167.2010.02598.x

17. Vovk T, Jakovljević MB, Kos MK, Janković SM, Mrhar A, Grabnar I. A Nonlinear Mixed Effects Modelling Analysis of Topiramate Pharmacokinetics in Patients with Epilepsy. Biological and Pharmaceutical Bulletin. 2010;33(7):1176–1182. doi:10.1248/bpb.33.1176

18. Jovanović M, Sokić D, Grabnar I, et al. Population pharmacokinetics of topiramate in adult patients with epilepsy using nonlinear mixed effects modelling. European Journal of Pharmaceutical Sciences. 2013;50(3):282–289. doi:10.1016/j.ejps.2013.07.008

19. Bae EK, Lee J, Shin JW, et al. Factors influencing topiramate clearance in adult patients with epilepsy: A population pharmacokinetic analysis. Seizure. 2016;37:8–12. doi:10.1016/j.seizure.2016.02.002

20. Takeuchi M, Yano I, Ito S, et al. Population Pharmacokinetics of Topiramate in Japanese Pediatric and Adult Patients With Epilepsy Using Routinely Monitored Data. Therapeutic Drug Monitoring. 2017;39(2):124. doi:10.1097/FTD.0000000000000383

21. Ahmed GF, Marino SE, Brundage RC, et al. Pharmacokinetic–pharmacodynamic modelling of intravenous and oral topiramate and its effect on phonemic fluency in adult healthy volunteers. British Journal of Clinical Pharmacology. 2015;79(5):820–830. doi:10.1111/bcp.12556

22. 22. Clark AM. Development of Intravenous Topiramate for Neuroprotection and Seizure Control in Neonates.

23. Cockcroft DW, Gault MH. Prediction of creatinine clearance from serum creatinine. Nephron. 1976;16(1):31–41. doi:10.1159/000180580

24. Tippayachai P, Leelakanok N, Methaneethorn J. Significant predictors for topiramate pharmacokinetics: a systematic review of population pharmacokinetic studies. Journal of Pharmacy Practice and Research. 2022;52(2):94–107. doi:10.1002/jppr.1787

25. Wei S, Li X, Zhang Q, et al. Population pharmacokinetics of topiramate in Chinese children with epilepsy. Eur J Clin Pharmacol. 2023;79(10):1401–1415. doi:10.1007/s00228-023-03549-6

